# Circular RNA and its mechanisms in diabetic retinopathy: a systematic review

**DOI:** 10.1101/2020.02.07.20021204

**Authors:** Miao He, Rouxi Zhou, Sen Liu, Weijing Cheng, Wei Wang

## Abstract

Circular RNAs (CircRNAs) are endogenous long non-coding RNAs. Unlike linear RNAs, they are structurally continuous and covalently closed, without 5 ’caps or 3’ polyadenylation tails. High-throughput RNA sequencing has enabled people to find several endogenous circRNAs in different species and tissues. circRNA mainly acts as a sponge for microRNAs in cytoplasm to regulates protein translation, or interacts with RNA-binding proteins to generate RNA protein complexes that control transcription. circRNAs are closely associated with diseases such as diabetes, neurological disorders, cardiovascular diseases and cancer, which indicates that circRNAs are closely related to and also play an important functional role in the occurrence and development of human diseases. Recent studies have shown that they are differentially expressed in healthy and diseased eye tissues. There lacks of biomarkers for early detection of diabetic retinopathy, and the newly discovered circRNAs seem to be an ideal candidate of novel molecular markers and therapeutic targets. However, the molecular mechanism of circRNAs activity in the occurrence and development of diabetic retinopathy are not clear yet. This systematic review aims to summarize the research status on function and mechanism of circRNAs in regulating the occurrence of diabetic retinopathy.

## 1. Introduction

Diabetic retinopathy (DR) is a major complication of diabetes, and it remains the leading cause of vision loss and blindness worldwide.^1, 2^ The pathogenesis of DR has been extensively studied for many years, mainly involving retinal microvascular dysfunction (including retinal nonperfusion, vascular permeability and retinal neovascularization) and neurodegeneration (including neuronal apoptosis and glial dysfunction).^3^ Clinically, based on retinal changes, DR can be classified into non-proliferative DR, which, under most circumstances, does not cause severe visual impairment, and proliferative retinopathy which can lead to significant visual loss or even blindness. Currently, methods for DR treatment include vitreous surgery, laser photocoagulation, and anti-VEGF drugs. However, the prognosis for DR patients remains poor, especially for those with advanced PDR. Therefore, it is necessary to develop new diagnostic and therapeutic technologies based on the pathogenesis of DR.

Circular RNAs (circRNAs) are a new type of RNA molecule characterized by covalently closed circle and widely present in eukaryotes.^4, 5^ They are derived from the exon or intron region of a gene, and are abundant in mammalian cells. Existing studies have shown that most circRNAs are conserved among different species. At the same time, they are stable because their circular structure can resist degradation of RNase R. CircRNA has attracted more and more attention due to its specificity of expression and complexity of regulation, and its important role in the occurrence of diseases, and has become a new research hotspot in the field of RNA.^6-8^

## 2. Research events of circRNA

The research process of circRNA has gone through three stages: discovery period, development period and outbreak period.^9^ In 1976, circular RNA molecules were observed in the cytoplasm of eukaryotic cells under an electron microscope. In 1993, circular transcripts were found in Sry gene of mice. In 1995, circRNA was found to achieve rolling circle translation in eukaryotes. In 2006, lariat-derived circRNA was discovered in RNase R digests. In 2010, a sponge model of non-coding RNA was proposed. In 2012, based on high-throughput sequencing technology, it was discovered that there existed a large amount of circRNAs in eukaryotes. In 2013, two articles about circRNA were published in the same issue of “Nature”.^7, 8^ Since then, circRNA related research has grown rapidly and gradually became a new star molecule in the field of RNA.Compared to linear RNA, the main features of circRNA include: (1) circRNA is produced by special alternative splicing, with abundant expression. (2) It’s expression is specific in terms of species, tissue and time. (3) circRNA has closed circular structure, which is not easily degraded by exonuclease and is more stable than linear RNA. (4) It is evolutionarily conserved across species. (5) It regulate gene expression at transcriptional and post-transcriptional levels. (6) Most circRNAs are non-coding, but a few can be translated into peptides.^10-12^

## 3. Biosynthesis of circRNA

The circRNA can derive from almost any part of the genome (exons and non-coding, transcripts are antisensed to 5 ’and 3’utrs or intergenic regions), which results in significant differences in molecular length. CircRNA can mainly be divided into three types according to its source: exon-derived circRNAs (exonic circRNAs), intron-derived circRNAs (circular intronic RNAs, ciRNAs) and circRNAs consisting of exons and introns (retained-intron circRNAs).^4, 13^ Figure 1 shows the main types of circRNA according to their source. Most exonic circRNAs are found in the cytoplasm, while the other two types are mainly in the nucleus.

**Figure 1.**
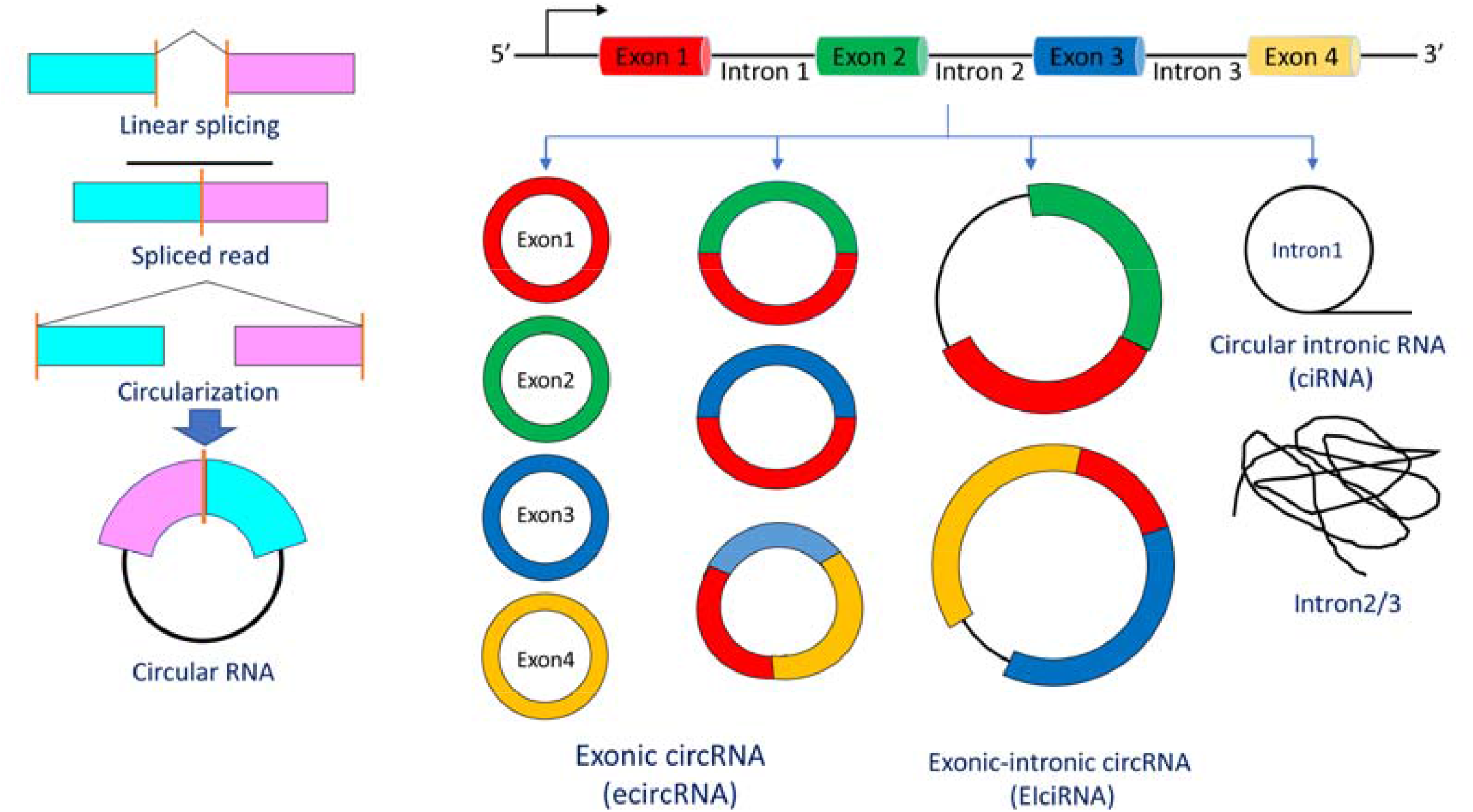
Classifications of circular RNA.

The circRNA is synthethized by “back-splicing” mechanism, which competes with and is significantly different from canonical splicing from which linear RNA molecules are generated. circRNA is produced by special pre-mRNA through alternative splicing, and its generation can be summarized into two categories: exon circularization and intron circularization. exonic circRNA mainly exists in the cytoplasm and is produced by pre-mRNA back-splicing, in which the splice donor (SD) of the downstream exon is connected to the splice acceptor (SA) of the upstream exon. And its formation mechanism can be mainly divided into two categories: direct back-splicing and exon skipping. Intron circularization mainly occurs in the nucleus. Its formation relies on nucleotide GU-rich sequences close to the 5′ splice site and nucleotide C-rich sequences near the branch point, with a small number of miRNA targets. Currently, there are mainly 5 models of circRNA formation: (1) exon skipping, (2) direct back-splicing, (3) intron pairing-driven circularization, (4) RBPs-dependent circularization, (5) alternative circularization mode similar to alternative splicing.^10^

The circRNA expression has the following unique characteristics: (1) Various alternative splicing patterns: a junction point may correspond to multiple alternative splicing patterns. (2) Presence of dominant circRNA: There are many types of circRNAs from the same gene, but they are usually dominated by a few types (dominant pattern). (3) circRNA and mRNA are disproportionate: The expression abundance of some circRNAs is related to the host gene, but that of most of them is not strongly related. (4) Tissue and disease specificity: The dominant circRNA pattern and alternative splicing method of the same gene differ in different tissues / diseases. (5) circRNA has a high overall expression abundance. (6) circRNA expression association in different tissues: circRNA expression overlaps and differs in different tissues / diseases.

The influencing factors of circRNA generation and the mechanism of tissue / disease-specific expression are not fully understood. At present, studies have found that the formation of circular RNA is regulated by multiple factors, including combined action of cis-regulatory element (CRE) and trans-splicing factors, including heterogeneous ribonucleic acid proteins (hnRNPs) and SR (Ser/Arg) proteins (proteins containing long repeating serine and arginine amino acid residues). There are some other proteins and molecules involved in the regulation of biosynthesis of circular RNA. For example, ADAR (double-stranded RNA (dsRNA) -specific adenosine deaminase that prevents the activation of the innate immune system) combine with double-stranded RNA and edits adenosine to inosine. And ATP-dependent RNA helicase A (also known as DHX9) inhibits the biogenesis of circular RNA by reversely repeating base-pairing between each other. There are also some proteins or molecules that can promote the formation of circular RNA, such as long flanking introns, inverted repeat elements (such as Alu element) and trans RNA-binding proteins (RBPs), RNA-binding protein FUS, protein quaking (HQK), NF90 and NF110, all of which are the protein products of the interleukin enhancer binding factor 3 gene and contribute to back-splicing.^14^

In addition, epigenetic changes in histones and genomes can affect alternative splicing and may also directly affect the biogenesis of circular RNA. For example, a recent study has shown that knockdown of DNMT3B, which is a key methyltransferase necessary for the maintenance of DNA methylation, can change the circular RNA expression, while these changes are not related to changes in the expression of the corresponding linear host genes. The degradation mechanism of circRNA remains elusive, which might be mediated by RNAi mechanism, circRNA degradation mediated by m6A modification, circRNA degradation by RNase L after viral infection, circRNA degradation mediated by TNRC6A/B/C. Recent study also suggested that m6A modification of CircRNA was tissue-specific expression, mediated circRNA degradation, and regulated nuclear export of circRNA.

## 4. Function of CircRNAs

The circRNAs have various functions, which are as follows (Figure 2):

**Figure 2.**
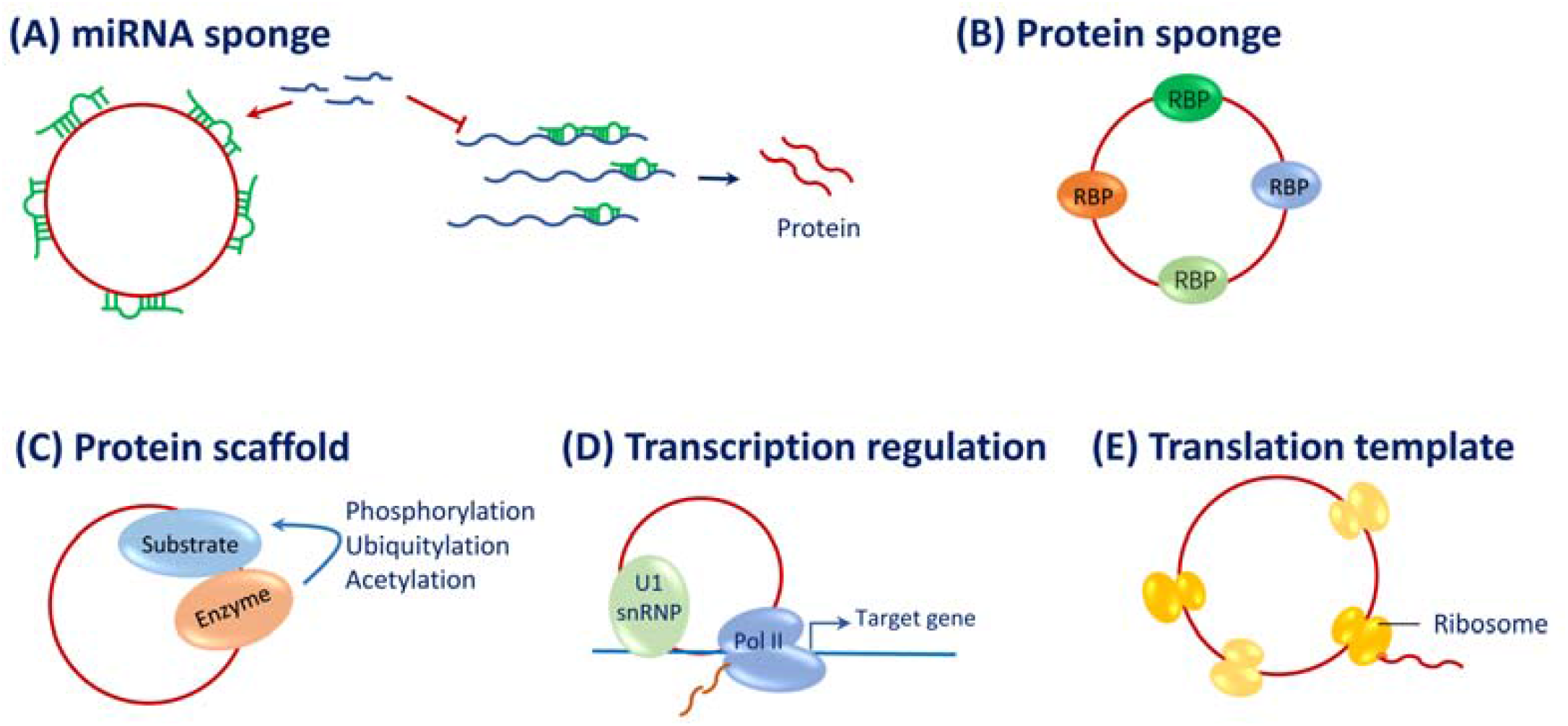
Main biological functions of circular RNA.

### 4.1 Competitively binds to miRNA (miRNA sponge)

Recent studies have shown that circular RNA molecules contain abundant microRNA (miRNA) binding sites and act as miRNA sponges in cells. It competitively binds to miRNA through miRNA response elements (MRE) to release the inhibitory effect of miRNA on its target genes and increase the expression level of the target genes. This mechanism of action is called the competitive endogenous RNA (ceRNA) mechanism. Studies have also shown that highly abundant circRNAs that contain many competing binding sites are more likely to have competitive endogenous RNA functions. For example, in many tissues, especially in the brain, there exists ciRS-7, which has a highly stable expression, that contains more than 70 miR-7 binding sites, and achieves competitive adsorption of miR-7 through the AGO2 protein to regulate target genes.^8^

### 4.2 Competitively binds to protein (protein sponge)

The circRNA can also regulate protein function by binding to RNA-binding proteins, such as inhibiting gene transcription by binding to transcription factors. circRNA can directly bind to proteins or indirectly associate with proteins mediated byRNA, and affect functions of the protein.It has been reported that a variety of circRNAs can bind to protein factors such as RNA-pol [ and AGO to act on the transcription initiation region, so as to regulate transcription. Ashwal-Fluss et al. found that the RNA splicing factor MBL could bind to exon 2 of its parent gene and promote its productionof circMbl, while circMbl could also bind to MBL to inhibit its activity and reduce the production of circMbl.

### 4.3 Acts as a linker or scaffold molecule for protein complexes (protein scaffolding)

In 2016, a study showed for the first time that circANRILl could be used as a protein scaffolding. In NIH3T3 mouse fibroblasts, circFOXO3 was found to interact with p21 and CDK2, respectively. And the formation of the circFOXO3-p21-CDK2 ternary complexes hindered the function of CDK2 and subsequently inhibited the cell cycle progress.

### 4.4 Being translated directly into protein or peptide (encoding protein)

Although circRNAs are non-coding RNAs, there are also some circRNAs that can encode polypeptides and exercise regulatory functions through the polypeptides.

### 4.5 Others

The circRNA was reported to regulate the process of transcription. The circRNA was also reported to have functions related to high-level structures, such as ribozyme and prion-like activity etc.

## 5. Dysregulated circRNA in DR

Changes in microvascular circulation in the retina under hyperglycemia conditions include: proliferation of endothelial cells and increased permeability, abnormal neovascularization and edema. An elevated blood glucose level leads to oxidative stress, inflammation, neuronal dysfunction, retinal ganglion cell apoptosis and glial cell activation. Studies have shown that circRNAs are involved in the occurrence and development of vascular endothelial dysfunction and has a potential regulatory effect on DR. circRNA is an emerging research hotspot, but its role in DR diseases has been relatively less reported.

### 5.1 circRNA in DR patients and high-glucose induced HRVEC

Only a few studies have evaluated the dysregulated expression of circRNA in DR patients, animal models, or cell models (Table 1).^15-18^ Gu et al.^18^ reported that 30 circRNAs were more than two-fold higher in serum of T2DR patients than those in T2DM patients and the control group, and they verified 7 differentially expressed circRNAs (hsa_circRNA_063981, hsa_circRNA_404457, hsa_circRNA_100750, hsa_circRNA_406918, hsa_circRNA_104387, hsa_circRNA_103410 and hsa_circRNA_100192) by q-PCR. We identified 122 upregulated and 9 downregulated circRNAs in vitreous humour samples of PDR patients compared to control.^19^ Zhang et al.^15^ determined that 529 circRNAs were differentially expressed between diabetic and non-diabetic retinas. The circ_0005015 was up-regulated in the plasma, vitreous samples, and fibrovascular membranes of patients with diabetes, which regulates retinal endothelial cell proliferation, migration, and tube formation. Yao et al.^20^ discovered 91 abnormally expressed circRNAs in the preretinal membrane of patients with PVR, which affected the proliferation, migration and secretion of RPE cells.

**Table 1.** Studies evaluating circular RNA changes in diabetic retinopathy.

| Author (Year) | Samples | Method | DE | Related circRNA | Gene | Mechanism |
| --- | --- | --- | --- | --- | --- | --- |
| Liu (2019) | Mice | Microarray | 844 | has_circ_0074837<br>mmu_circ_0000254 | PWWP2A | miRNA sponge<br>miR-579 |
| Zhu (2019) | Rat | qRT-PCR | - | circDNMT3B | DNMT3B | miRNA sponge<br>miR-20b-5p |
| Yao (2019) | Human | Microarray | 91 | hsa_circ_0043144 | CCL3 | - |
| He (2019) | Human | RNA-seq | 131 | hsa_circ_0005941 | FTO | - |
| Shan (2017) | Mice | Microarray | - | has_circ_0000284<br>mmu_circ_0001052 | HIPK3 | miRNA sponge<br>miR-30a-3p |
| Liu (2017) | HUVEC | qRT-PCR | - | has_circ_0000615<br>mmu_circ_0000498 | ZNF609 | miRNA sponge<br>miR-615-5p |
| Zhang (2017) | Human | Microarray | 529 | hsa_circ_0005015 | HAS2 | miRNA sponge<br>miR-519d-3p |
| Gu (2017) | Human | Microarray | 30 | - | - | - |

DE=differential expression; HUVEC=human umbilical vein endothelial cell.

The currently identified circRNA play the role by miRNA sponge mechanism (Figure 3). The circ_0005015 acts as a sponge for miR-519d-3p, which inhibits its activity and interferes expressions of MMP-2, XIAP and STAT3.^15^ Shan et al.^21^ found that circHIPK3 in HRVECs was significantly up-regulated under high glucose stimulation. circHIPK3 acts as a sponge for endogenous miR-30a-3p, so as to inhibit its activity and cause increased expressions of vascular endothelial growth factor-C (VEGF-C), wingless-type mouse mammary tumor virus integration site family member 2 (WNT2) and frizzled-4 (FZD4), which prompts that circHIPK3 causes vascular endothelial proliferation dysfunction by blocking miR-30a. Liu et al.^22^ revealed that diabetic stress up-regulates the expression of cPWWP2A in periretinal cells. CPWWP2A acts as a sponge for endogenous miR-579 and regulates the expression of Angiopoietin 1 / Occludin / SIRT1 genes, which directly regulates the biological functions of pericytes. Meanwhile, exosomes carrying cPWWP2A indirectly regulate the biological functions of ECs, so as to promote ECs migration and angiogenesis. Zhu et al.^23^ found that the circDNMT3B gene expression in the retinal tissue of DR patients was down-regulated, while the miR-20b-5p gene was up-regulated, and they verified these results in diabetic animal models.

**Figure 3.**
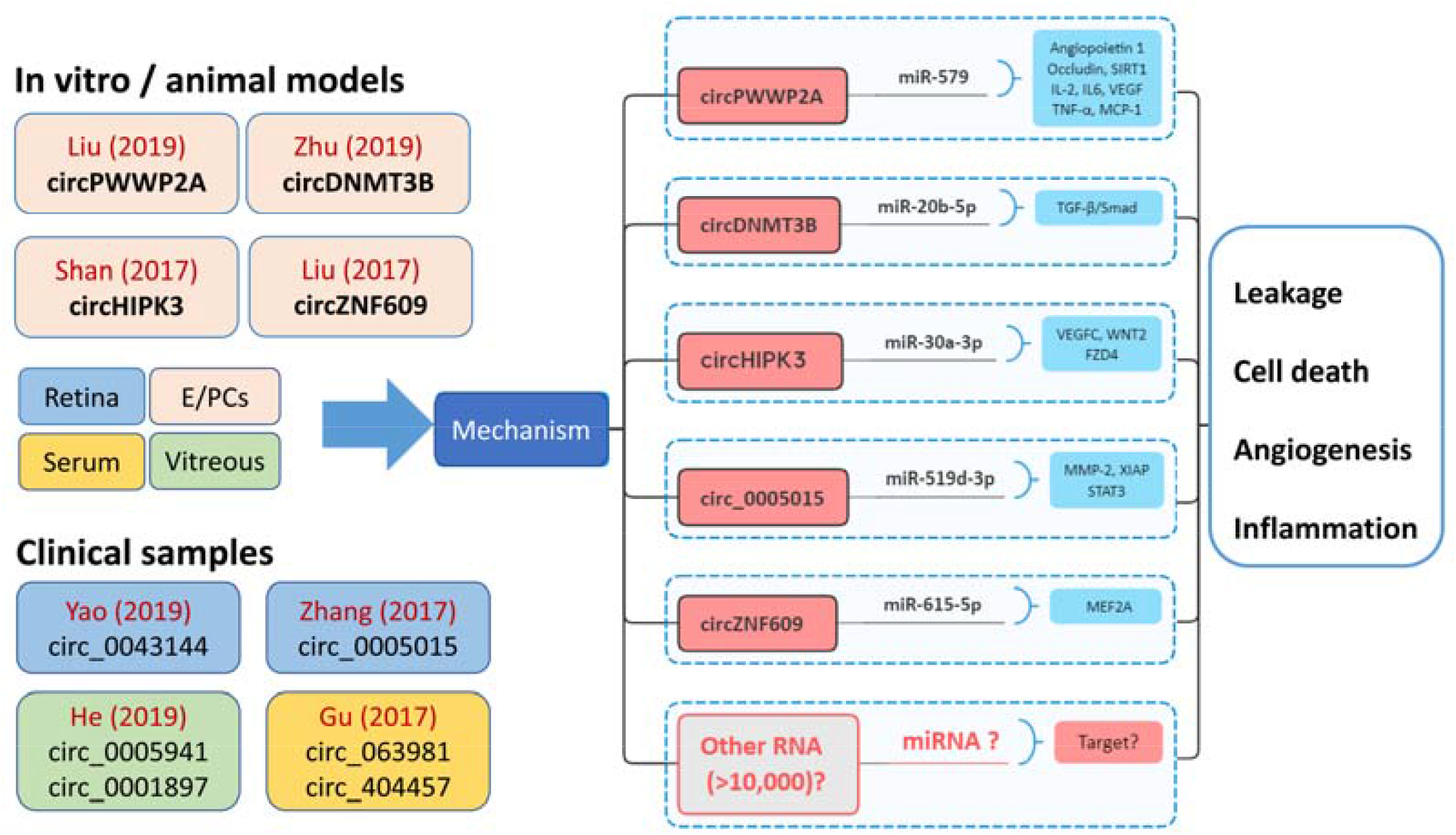
Main finding of dysregulated circular RNA and DR studies.

Down-regulation of circDNMT3B caused by hyperglycemia induces abnormal retinal microvascular function by regulating miR-20b-5p / BAMBI. Meanwhile, overexpression of circDNMT3B gene can reduce retinal vascular function damage caused by diabetes and significantly improve visual function.

### 5.2 circRNA in endothelial cells of macro-vasculature

Vascular endothelial cell dysfunction is the initiating step for the occurrence and development of DR and plays a decisive role in the prognosis of DR. Besides from HRVEC, the human umbilical vein endothelial cells (HUVEC) and human aortic endothelial cells (HAEC) were commonly used. Shang et al.^24^ reported that more than 1,000 new circRNAs and 95 differentially expressed circRNAs were detected in HUVEC cultured in high glucose environment in vitro. Jin et al.^25^ identified 214 differentially expressed circRNAs in high glucose induced HUVEC compared with normal controls by using RNA sequencing, of which 7 were validated using qPCR (hsa_circ_0008360, hsa_circ_0005741, hsa_circ_0003250, hsa_circ_0045462, hsa_circ_0064772, hsa_circ_0007976 and hsa_circ_0005263). Huang et al.^26^reported that in rat HAEC induced by transforming growth factor β1 (TGF-β1), a total of 102 circRNA was differentially expressed, of which 66 circRNA expressions were up-regulated and 26 circRNA expressions were down-regulated.

Angiogenesis: In 2015, a study for the first time reported that circRNAs were expressed in endothelial cells and were regulated by hypoxia in vitro.^27^ Boeckel et al.^28^ found that the expressions of circZNF292, circAFF1 and circDENND4C were increased and that of circTHSD1 was decreased in HUVEC under hypoxia. Among them, the silencing of cZNF292 significantly inhibited the proliferation of endothelial cells and the formation of vascular tube, and the increased level of cZNF292 by transfection plasmid could increase the proliferation of endothelial cells. Pan et al.^29^ reported that the expression of circ_0054633 in HUVEC increased in a high glucose (HG) environment, while down-regulation of circRNA-0054633 increased high glucose-induced endothelial cell dysfunction and angiogenesis. Bioinformatics analysis has shown that circRNA-0054633 may inhibit miR-218 expression, which further lead to the upregulation of roundabout1 (ROBO1) and heme oxygenase-1 (HO-1), and as a result, high glucose-induced endothelial cell dysfunction was inhibited. Liu et al.^30^ reported that circZNF609 was significantly up-regulated in HUVEC induced by high glucose and hypoxia, and silencing circZNF609 could inhibit the migration of vascular endothelial cells and tube formation, protect endothelial cells and enhance their resistance against oxidative stress injury and hypoxic stress injury. Mechanism studies have revealed that circZNF609 up-regulates the expression of myocyte enhancer factor 2A (MEF2A) by inhibiting the activity of miR-615-5p through the sponge mechanism, and then exerts biological functions that regulate angiogenesis.

Apoptosis: Dang et al.^31^ found that in HUVEC exposed to hypoxia, has_circ_0010729 expression was significantly up-regulated, while knockout of has_circ_0010729 could inhibit cell proliferation, and promote apoptosis. Cao et al.^32^ found that circHIPK3 was down-regulated in HUVEC and HAEC of diabetic patients after HG treatment. In HUVEC and HAEC, lentivirus-mediated overexpression of circHIPK3 inhibited HG-induced cell death and apoptosis, which suggests that down-regulation of circHIPK3 mediates HG-induced endothelial cell injury.

Inflammation: Cheng et al.^33^ found that the down-regulation of hsa_circ_0068087 could alleviate the TLR4 / NF-κB / NLRP3 inflammasome-mediated inflammation and endothelial cell dysfunction by acting as a miR-197 sponge in high-glucose induced HUVEC.

## 6. Conclusion

The study of circRNA has brought us many surprising discoveries, which means that circRNA is of important significance biologically and pathologically. circRNA plays an important role in DR by regulating the angiogenesis, proliferation, apoptosis and inflammatory response of various cells in the retina. Hence, the exploration of the role of circRNA in DR will help provide new ideas and solutions for the prevention and control of DR development and progression. circRNA has great potential as an effective regulatory molecule. Due to its cell type and disease-specific expression, circRNA can not only serve as a DR biomarker, but also become a therapeutic target for DR. Although there is conclusive evidence that circRNA is involved in the development of DR, the extensiveness and complexity of transcription of circRNA is seriously underestimated, and the role of most circRNAs in DR and their effective use as a therapeutic molecule are subject to further research.

## 7. Methods of Literature Search

The electronic databases including Pubmed (http://www.ncbi.nlm.nih.gov/pubmed) and Embase.com (http://www.embase.com) were systematic searched from the inception to January 2020. The ARVO Meeting Abstracts website (http://www.iovs.org/search.dtl?arvomtgsearch=true) was retrieved for additional study. The following key words were used: circular RNA, non-coding RNA, circRNA, diabetic retinopathy, diabetic eye diseases, diabetic ocular complications, endothelial cell. All abstracts were reviewed and appropriate studies were obtained in full for complete review and inclusion. The references of included studies and related reviews were hand-screened for additional articles. When multiple articles involving same subjects were available, the most comprehensive one was adopted. The articles in English were included, and those in languages other than English were considered when English abstracts were available.

## Data Availability

This is a systemic review, all data referred to in the manuscript is available.

## Acknowledgments

The authors declare no conflict of interest in respect to the topic and content of this manuscript. This study was supported by the National Natural Science Foundation of China (81900866).

## References

1. Zheng Y, Ley SH, Hu FB. Global aetiology and epidemiology of type 2 diabetes mellitus and its complications. Nat Rev Endocrinol 2018;14:88–98.

2. Leasher JL, Bourne RR, Flaxman SR, et al. Global Estimates on the Number of People Blind or Visually Impaired by Diabetic Retinopathy: A Meta-analysis From 1990 to 2010. Diabetes Care 2016;39:1643–1649.

3. Stitt AW, Curtis TM, Chen M, et al. The progress in understanding and treatment of diabetic retinopathy. Prog Retin Eye Res 2016;51:156–186.

4. Kristensen LS, Andersen MS, Stagsted L, Ebbesen KK, Hansen TB, Kjems J. The biogenesis, biology and characterization of circular RNAs. Nat Rev Genet 2019.

5. Thomson DW, Dinger ME. Endogenous microRNA sponges: Evidence and controversy. Nat Rev Genet 2016;17:272–283.

6. Tay Y, Rinn J, Pandolfi PP. The multilayered complexity of ceRNA crosstalk and competition. Nature 2014;505:344–352.

7. Memczak S, Jens M, Elefsinioti A, et al. Circular RNAs are a large class of animal RNAs with regulatory potency. Nature 2013;495:333–338.

8. Hansen TB, Jensen TI, Clausen BH, et al. Natural RNA circles function as efficient microRNA sponges. Nature 2013;495:384–388.

9. Patop IL, Wust S, Kadener S. Past, present, and future of circRNAs. Embo J 2019;38:e100836.

10. Aufiero S, Reckman YJ, Pinto YM, Creemers EE. Circular RNAs open a new chapter in cardiovascular biology. Nat Rev Cardiol 2019;16:503–514.

11. Li X, Yang L, Chen LL. The biogenesis, functions, and challenges of circular RNAs. Mol Cell 2018;71:428–442.

12. Salzman J, Chen RE, Olsen MN, Wang PL, Brown PO. Cell-type specific features of circular RNA expression. Plos Genet 2013;9:e1003777.

13. Ebbesen KK, Kjems J, Hansen TB. Circular RNAs: Identification, biogenesis and function. Biochim Biophys Acta 2016;1859:163–168.

14. Pagliarini V, Jolly A, Bielli P, Di Rosa V, De la Grange P, Sette C. Sam68 binds Alu-rich introns in SMN and promotes pre-mRNA circularization. Nucleic Acids Res 2019.

15. Zhang SJ, Chen X, Li CP, et al. Identification and characterization of circular RNAs as a new class of putative biomarkers in diabetes retinopathy. Invest Ophthalmol Vis Sci 2017;58:6500–6509.

16. Cao M, Zhang L, Wang JH, et al. Identifying circRNA-associated-ceRNA networks in retinal neovascularization in mice. Int J Med Sci 2019;16:1356–1365.

17. Sun LF, Zhang B, Chen XJ, et al. Circular RNAs in human and vertebrate neural retinas. Rna Biol 2019.

18. Gu Y, Ke G, Wang L, Zhou E, Zhu K, Wei Y. Altered expression profile of circular RNAs in the serum of patients with diabetic retinopathy revealed by microarray. Ophthalmic Res 2017;58:176–184.

19. He M, Wang W, Yu H, et al. Comparison of expression profiling of circular RNAs in vitreous humour between diabetic retinopathy and non-diabetes mellitus patients. Acta Diabetol 2019.

20. Yao J, Hu LL, Li XM, et al. Comprehensive circular RNA profiling of proliferative vitreoretinopathy and its clinical significance. Biomed Pharmacother 2019;111:548–554.

21. Shan K, Liu C, Liu BH, et al. Circular noncoding RNA HIPK3 mediates retinal vascular dysfunction in diabetes mellitus. Circulation 2017;136:1629–1642.

22. Liu C, Ge HM, Liu BH, et al. Targeting pericyte-endothelial cell crosstalk by circular RNA-cPWWP2A inhibition aggravates diabetes-induced microvascular dysfunction. Proc Natl Acad Sci U S A 2019;116:7455–7464.

23. Zhu K, Hu X, Chen H, et al. Downregulation of circRNA DMNT3B contributes to diabetic retinal vascular dysfunction through targeting miR-20b-5p and BAMBI. Ebiomedicine 2019.

24. Shang FF, Luo S, Liang X, Xia Y. Alterations of circular RNAs in hyperglycemic human endothelial cells. Biochem Biophys Res Commun 2018;499:551–555.

25. Jin G, Wang Q, Hu X, et al. Profiling and functional analysis of differentially expressed circular RNAs in high glucose-induced human umbilical vein endothelial cells. Febs Open Bio 2019;9:1640–1651.

26. Huang X, Chen Y, Xiao J, et al. Identification of differentially expressed circular RNAs during TGF-ss1-induced endothelial-to-mesenchymal transition in rat coronary artery endothelial cells. Anatol J Cardiol 2018;19:192–197.

27. Memczak S, Papavasileiou P, Peters O, Rajewsky N. Identification and characterization of circular RNAs as a new class of putative biomarkers in human blood. Plos One 2015;10:e141214.

28. Boeckel JN, Jae N, Heumuller AW, et al. Identification and characterization of Hypoxia-Regulated endothelial circular RNA. Circ Res 2015;117:884–890.

29. Pan L, Lian W, Zhang X, et al. Human circular RNA0054633 regulates high glucoseinduced vascular endothelial cell dysfunction through the microRNA218/roundabout 1 and microRNA218/heme oxygenase1 axes. Int J Mol Med 2018;42:597–606.

30. Liu C, Yao MD, Li CP, et al. Silencing of circular RNA-ZNF609 ameliorates vascular endothelial dysfunction. Theranostics 2017;7:2863–2877.

31. Dang RY, Liu FL, Li Y. Circular RNA hsa_circ_0010729 regulates vascular endothelial cell proliferation and apoptosis by targeting the miR-186/HIF-1alpha axis. Biochem Biophys Res Commun 2017;490:104–110.

32. Cao Y, Yuan G, Zhang Y, Lu R. High glucose-induced circHIPK3 downregulation mediates endothelial cell injury. Biochem Biophys Res Commun 2018;507:362–368.

33. Cheng J, Liu Q, Hu N, et al. Downregulation of hsa_circ_0068087 ameliorates TLR4/NF-kappaB/NLRP3 inflammasome-mediated inflammation and endothelial cell dysfunction in high glucose conditioned by sponging miR-197. Gene 2019;709:1–7.

